# The impact of a home-based personalized computerized training program on cognitive dysfunction associated with Long COVID: a before-and-after feasibility study

**DOI:** 10.1101/2022.09.28.22280467

**Authors:** Jon Andoni Duñabeitia, Francisco Mera, Óscar Baro, Tamen Jadad-Garcia, Alejandro R. Jadad

## Abstract

**Background:** Long COVID—also known as post-acute sequelae of COVID-19 or PASC—is a systemic syndrome affecting a large number of persons in the aftermath of the pandemic. Cognitive dysfunction (or brain fog) is one of its most common manifestations of PACS, and there are no effective interventions to mitigate it. Home-based personalized computerized cognitive training (CCT), which has shown effectiveness to improve other conditions, could offer hope to relieve the cognitive dysfunction in people with a previous history of COVID-19.

**Objective:** To evaluate the feasibility and potential benefit of a personalized CCT intervention to improve cognitive function among people living with PACS.

**Methods:** Adult individuals who self-reported cognitive dysfunction more than 3 months after a diagnosis of COVID-19 were recruited through an online platform designed for the study. Those who were eligible assessed their general cognitive function before completing as many cognitive daily training sessions as they wished during an 8-week period, using a personalized CCT application at home. The sessions included gamified tasks that tapped into five cognitive domains (attention, coordination, memory, perception and reasoning) and were tailored to the specific cognitive strengths and weaknesses detected at each point. At the end of this period, participants repeated the general cognitive function assessment. The differences between the scores at 8 weeks and baseline was the main outcome, complemented with analyses of the changes based on the participants’ age, training time, self-reported health level at baseline and time since the initial COVID-19 infection. Participants’ cognitive assessment scores were also computed in terms of percentiles according to the normative database of the test, considering their corresponding age- and gender-based reference sample.

**Results:** The participants had significant cognitive dysfunction at baseline, even though 80% of them had had the initial episode of COVID-19 more than a year before enrolling in the study. Eighty nine percent reported negative levels of self-reported health at baseline. On average, 51 training sessions (range: 10 to 251) were completed over a mean time of 435 minutes (range 78 to 2448). Most of the participants obtained higher scores after CCT in each of the domains as compared with baseline (attention: 81% of the sample; memory: 86%; coordination: 82%; perception: 88%; reasoning: 77%). The magnitude of the score increase at post-test was high across domains (attention: 31% of change; memory: 37%; coordination: 52%; perception: 42%; reasoning: 26%). Following CCT, there were also improvements in the percentile data in all the domains (attention: 14 points; memory: 18 points; coordination: 18 points; perception: 17 points; reasoning: 11 points).

**Conclusions:** This study suggests that a self-administered CCT based on gamified cognitive tasks could be an effective way to ameliorate cognitive dysfunction in persons with PASC.

## Introduction

The coronavirus disease 2019 (COVID-19) pandemic was caused by the pathogen known as Severe Acute Respiratory Syndrome Coronavirus 2 (SARS-CoV-2). Now understood to be a multi-organ illness with a wide range of symptoms, COVID-19 has also resulted in post-acute reports of symptoms and structural organic changes that are extended and chronic, bringing unprecedented levels of additional morbidity. These long-term impacts—known collectively as post-acute sequelae of COVID-19 (PASC) (Soriano et al., 2021)—range from minor complaints to major diseases, reaching even near-fatal situations in some cases, all having a direct impact not only in the physical but also in the psychological health of those affected, and in their levels of productivity (Davis et al., 2021; Reuschke & Houston, 2022).

PASC comprises a plethora of pulmonary, hematologic, cardiovascular, renal, endocrine, gastrointestinal, dermatologic, and neuropsychiatric and neuropsychological sequelae. Comprehensive large-scale cohort studies have revealed that COVID-19 survivors are at a significantly higher risk of developing neurologic problems than their non-infected peers beyond the first 30 days of infection (see Xu, Xie & Al-Aly, 2022). One of the most prevalent manifestations of PACS, which affects around one in three individuals, is generalized cognitive dysfunction (e.g., Deer et al., 2021; Premraj et al., 2022; Davis et al., 2021; Taquet et al., 2021), similar to the long-term consequences of prior epidemics and other infections (see Islam et al., 2020; Stefano, 2021).

Although a unified set of diagnostic criteria for PASC is still lacking, there is consensus across major organizations about the inclusion of dysfunctional cognition as a key component (e.g., Centers for Disease Control and Prevention, 2022; National Institute for Health and Care Excellence, 2021; World Health Organization, 2021). While the underlying causes driving this cognitive dysfunction in PASC are still unclear, research suggests that the persistence of viral infection and immune dysregulation could play a key role in its etiology (Moghimi et al., 2021).

Despite the knowledge gaps about its root causes, researchers are starting to explore multiple options to curb the consequences of PASC, drawing from studies in populations with similar signs and symptoms emanating from other causes (Ledford, 2022). Along these lines, an option that could be valuable for the management of cognitive dysfunction in people with PASC is personalized computerized cognitive training (CCT), an approach that includes the administration of gamified exercises through digital devices, typically at home. CCTs have been shown to yield significant improvements of dysfunctional cognitive abilities associated with stroke, Parkinson’s disease, age-related cognitive impairment or multiple sclerosis, among many other conditions, with an intervention duration between 4 and 12 weeks across conditions (Gavelin et al., 2022; Zhou et al., 2022; see also Bahar-Fuchs et al., 2020, Embon-Magal et al., 2022, and Gigler et al., 2013, for a successful 8-week intervention). In light of this, to the best of our knowledge, we describe what is the first effort to investigate the potential role of CCT to ameliorate cognitive dysfunction in people living with PASC.

## Methods

This was a single-center before-and-after study. The details are reported in line with the CONSORT extension to pilot and feasibility trials (Lancaster and Thabane, 2019).

The study protocol was approved by the Ethics Committee of Universidad Nebrija, Spain, following the criteria set by the Declaration of Helsinki. Personal data from all participants were treated confidentially according to the applicable General Data Protection Regulations of the European Union, which are deemed to be the toughest in the world. Records were anonymized and encrypted in secure servers to provide further data security.

### Settings, sampling frame and timelines

The recruitment of the candidate participants was led by the CIR Long COVID Research Center (https://cirlongcovid.org), an institution aimed to generating multidisciplinary high-resolution services for people affected by PASC, in collaboration with the Centro de Investigación Nebrija en Cognición (CINC) of Nebrija University (Madrid, Spain). The contact point of Long Covid ACTS (Autonomous Communities Together Spain), the main Spanish PASC patient association, disseminated an online message among its affiliates, explaining the study and inviting them to consider enrolling in it. Potential participants sent individual expressions of interest via an online platform specifically designed for this study (https://persistente.org/), which they could access from anyplace using any device connected to the Internet. Recruitment took place between October 2021 and December 2021, and participants that met the inclusion criteria were accepted on a rolling basis. The first participant completed the study in December 2021, and the last one in February 2022. All phases of the study were conducted online.

Once eligible participants had been accepted, they completed the cognitive assessments and training in an unsupervised manner. It should be noted in this regard that cognitive dysfunction (or ‘brain fog’) is a debilitating manifestation of PASC with different levels of severity that still allow individuals to complete tasks and activities (Bungenberg et al., 2022).

### Selection criteria and enrolment

Potentially eligible individuals were included in the study if they: 1) were adults (older than 18 years old), 2) reported having been infected with COVID-19 at least 3 months prior to their expression of interest, and 3) experienced cognitive symptoms associated with PACS (concentration problems or brain fog).

A trained neuroscientist from the research team (JAD) reviewed all self-nominations and identified the candidates who met the selection criteria, excluding those who did not meet them.

All participants who met the inclusion criteria signed an informed consent form prior to their enrollment in the study. At that point, their identity was validated by using a procedure based on confirmation emails required to access such a form.

### Materials and Procedure

Upon enrolment, included participants completed an online questionnaire with items designed to capture data on their sociodemographic information, their history of infection with COVID-19 and their health status. They were asked to rate their self-perceived estimated percentage of health loss with respect to time that immediately preceded the COVID-19 infection (options: <25%, 25%-50%, 50%-75%, >75%), as well as to indicate their self-reported level of health (*“In general, would you rate your health as excellent, very good, good, fair or poor?”*). Immediately after this they were asked to complete the Cognitive Assessment Battery (CAB)™ PRO (CogniFit Inc., San Francisco, US; https://www.cognifit.com/cab). The CAB is a self-administered online general cognitive evaluation psychometric tool (see Buades-Sitjar & Duñabeitia, 2022; Yaneva et al., 2022) that takes 30-35 minutes to complete using either a laptop, desktop or tablet computer. The CAB includes a series of 17 short tests that evaluate a variety of different cognitive abilities, putting a heavy focus on executive functions. These are then used to obtain a gender- and age-adjusted general score, which ranges from 0 to 800 points, as well as five different sub-scores based on the cognitive domains of perception, attention, memory, coordination and reasoning. The calculation of the cognitive score in each of the five domains is done by averaging the scores of the individual cognitive skills that constitute them (For reasoning: processing speed, shifting and planning; for memory: auditory short term memory, visual short term memory, short term memory, working memory, visual memory, contextualized memory, naming; for attention: inhibition, focus attention, updating, divided attention; for perception: visual scanning, auditory perception, estimation, recognition, visual perception, spatial perception; and for coordination: response time, eye-hand coordination). The z-score in each of the cognitive domains was obtained for each participant and used as the main outcome measure. These data were obtained using the reference normative dataset of the CAB, which was composed of 1,282,242 unique healthy test-takers (570,980 males and 711,262 females) as of September 2022.

Once the initial cognitive screening was completed, the digital platform automatically and consecutively assigned participants to the training phase. In this phase, participants were asked to complete short, gamified sessions of around 10 minutes each consisting of a variety of tasks specifically designed to tax and train the different cognitive skills. Each training session included two different gamified cognitive tasks selected from a pool of 12 activities. Each training program was tailored to the individuals’ specific cognitive strengths and weaknesses detected in the CAB by a patented Individualized Training System™ (ITS) software that automatically chooses the activities and difficulty levels for each person in every session. All individuals were asked to complete a training lasting for 8 weeks in which they could access the training platform as frequently as they desired. The data regarding the adherence of the participants to the training program (as measured by the total number of minutes invested in training sessions) were used as a secondary outcome measure.

After the 8 weeks of training, all participants completed the CAB again and new z-scores in each of the cognitive domains were calculated as compared to the normative sample.

### Data analysis

The difference in scores between the initial and final assessments was used as the main outcome measure. To this end, the z-transformed scores obtained by each participant in each of the five cognitive domains measured in the CAB before and after the CCT phase (attention, coordination, memory, perception, reasoning) were compared across test moments using both parametric (repeated measures ANOVA) and non-parametric tests (Kruskal-Wallis ANOVA) after exploring whether the data distribution departed from normality with the Shapiro-Wilk test. The ANOVA tests followed a 5 (Domain: attention, coordination, memory, perception, reasoning) by 2 (Test Moment: pre-test, post-test) design. In the presence of a significant interaction, post hoc pairwise comparisons were performed for each level of the Domain factor across the levels of Test Moment. A series of additional ANOVA tests explored the potential mediating role of participants’ age (measured in years; the Age variable) and the time devoted to the training (measured in minutes; the Number of Minutes of Training variable) using them as covariates. Additional analyses explored if their self-reported level of health at baseline (dichotomized into positive health for ‘good’, ‘very good’ or ‘excellent’; or negative for ‘poor’ or ‘fair’; and labeled as the Health at Baseline variable), and the time from COVID-19 infection (dichotomized into less than a year or more than a year from infection; and labeled as the Time from Infection variable) modulated the differences observed between pre- and post-test in each of the five cognitive domains using Fisher’s exact test calculation. Participants’ cognitive assessment scores were also computed in terms of percentiles according to the normative database of the test, and the mean values for each of the cognitive domains were obtained by considering each participant in relation with their corresponding age- and gender-based reference sample. Similarly, the participants’ gender- and age-corrected score in the 0-800 scale generated by the CAB was computed, where scores between 0 and 200 represent a cognitive performance that is well below average, scores between 201 and 400 correspond to a cognitive performance below average, and scores higher than 400 correspond to a cognitive performance above average. The data corresponding to the participants’ sociodemographic profile and their history of COVID-19 infection were analyzed using descriptive statistics. R-based *jamovi* statistical software was used to run the analysis (The jamovi project, 2021). In all cases, a p-value lower than 0.05 was considered statistically significant.

## Results

A total of 73 individuals (mean age = 46.1 years; SD = 7.6; 66 females) were included in the study. Two of the participants (2.7%) reported having been infected between 3 and 6 months before their enrolment in the study, 3 individuals (4.1%) between 6 and 9 months before, 10 individuals (13.7%) between 9 and 12 months before, 21 individuals (28.8%) between 12 and 18 months before and 37 individuals (50.7%) more than 18 months before. When asked about how they perceived their current overall health level as compared to the pre-infection stage, 45 individuals (61.6%) reported having lost at least 50% of their health level. Sixty five participants (89.0%) reported having poor or fair self-rated health at baseline.

The averaged age- and gender-corrected percentiles associated with the scores obtained in the CAB for the five investigated cognitive domains showed that the test sample was below the median value (50th percentile) across domains at pre-test, pointing to the existence of a generalized cognitive dysfunction (Table). Forty three (58.9%) of 73 participants scored below the 400 cut-off point in attention, 33 (45.2%) in memory, 61 (83.6%) in coordination, 53 (72.6%) in perception and 50 (68.5%) in reasoning.

The mean number of computerized cognitive training sessions completed across individuals was 51 (SD = 41; median = 44; range: 10-251) and the mean time invested in the intervention was 435 minutes (SD = 383; median = 358; range: 78-2448).

There was a consistent increase in the scores obtained in the cognitive assessment after the CCT (i.e., at post-test) as compared with those at baseline, and this increase extended to the five measured cognitive domains (Table). Only 25 (34.2%) out of 73 participants scored below the 400 cut-off point in attention, 18 (24.7%) in memory, 40 (54.8%) in coordination, 21 (28.8%) in perception and 29 (39.7%) in reasoning. The mean percentage of increase in the cognitive score as compared to baseline was 31% score increase for attention, 37% for memory, 52% for coordination, 42% for perception and 26% for reasoning.

When the percentile data calculated after CCT were contrasted with those at baseline, numerical improvements in all the domains were observed (attention: 14 percentile points; memory: 18 points; coordination: 18 points; perception: 17 points; reasoning: 11 points). Moreover, most of the participants obtained higher absolute scores after CCT in each of the domains, with 59 (81%) achieving improvements in attention, 63 (82%) in memory, 60 (82%) in coordination, 64 (88%) in perception, and 56 (77%) in reasoning scores.

The factorial analysis of the z-scores revealed a significant main effect of Test Moment (F(1,72) = 104.36, p < 0.001, η^2^ _partial_ = 0.592), suggesting that cognitive performance increased after training (mean difference = 0.63, t(72) = 10.2, Tukey-corrected p < .001). The interaction between the two factors was significant (F(4,288) = 9.48, p <0 .001, η^2^ _partial_ = 0.116), showing that the effect of the training varied as a function of the specific cognitive domain. Post hoc pairwise comparisons showed that scores in all domains improved with training (all ts > 4.25 and Tukey-corrected p-values < 0.01), but the mean differences revealed that the gains were not equal across domains (attention = 0.64, coordination = 1.01, memory = 0.54, perception = 0.62, reasoning = 0.36) (Figure 1).

**Figure 1.**
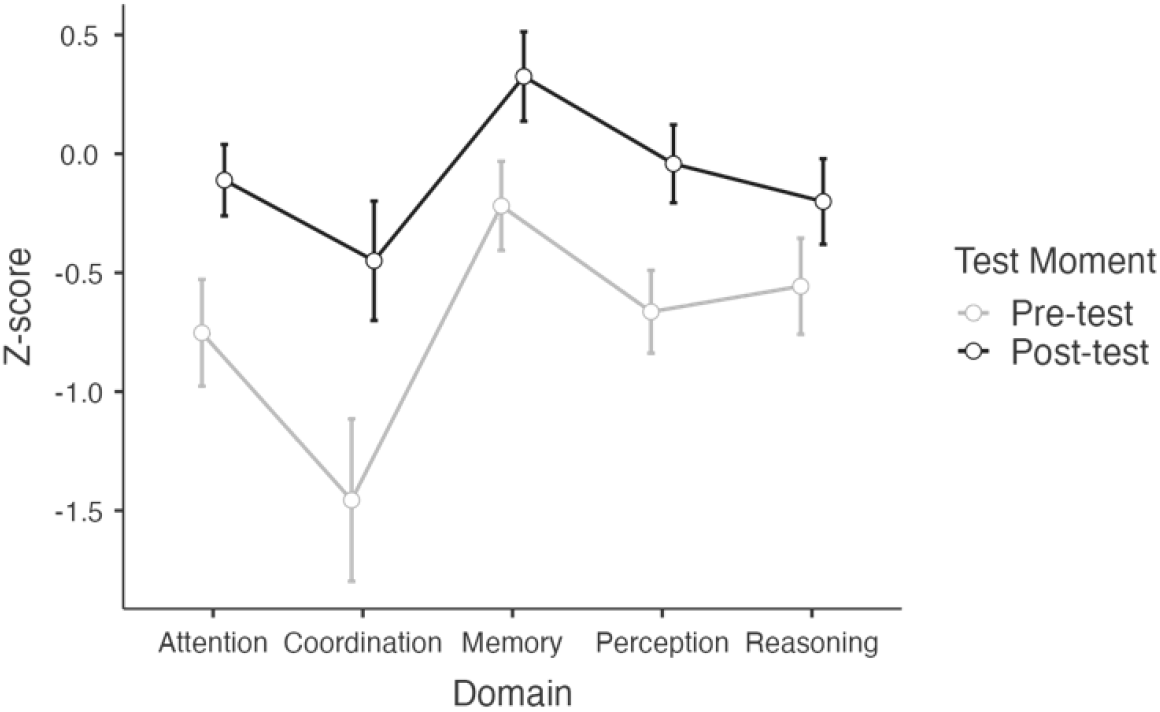
Factorial analysis of z-scores.

The analysis including the age of the participants and the number of minutes of training invested by each of them as covariates showed a significant interaction between the Test Moment and the Number of Minutes of Training (F(1,70) = 6.60, p = .012, η^2^ _partial_ = 0.086), indicating that the cognitive improvement increased as a function of the amount of training similarly for all domains (Figure 2). The main effects of Test Moment and Domain, and the interactions between these factors and Age were not statistically significant (all Fs < 1.88 and p-values > 0.11).

**Figure 2.**
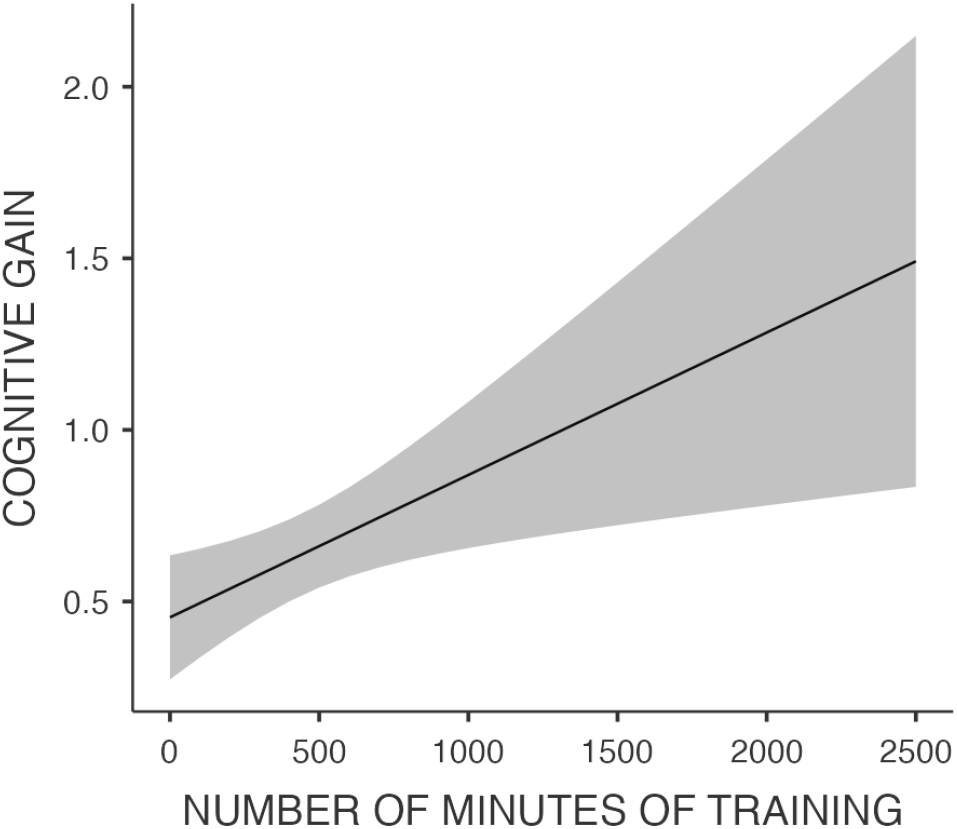
Relationship between minutes of training and cognitive gain (in z-scores)

The examination of the impact of the CCT in each individual cognitive domain as a function of participants’ level of self-reported health at baseline (positive vs. negative) and the time from COVID-19 infection (more vs. less than a year) showed that the existence of cognitive enhancement (presence vs. absence of improvements) did not depend on any of these variables (all p-values of the Fisher’s exact tests > 0.21).

Considering that the data were not normally distributed as evidenced by a series of Shapiro-Wilk normality tests (Ws between 0.896 and 0.986, with 8/10 tests being significant at the p < 0.05 level), a non-parametric Kruskal-Wallis ANOVA was run to validate the results. All the pre-test vs. post-test comparisons across Domains were significant (attention: χ^2^ = 16.11, p < 0.001, ε^2^ = 0.111; coordination: χ^2^ = 16.97, p < 0.001, ε^2^ = .117; memory: χ^2^ = 16.33, p < .001, ε^2^ = 0.113; perception: χ^2^ = 27.94, p < 0.001, ε^2^ = 0.193; reasoning: χ^2^ = 7.35, p < 0.01, ε^2^ = 0.051), endorsing the results of the parametric tests and demonstrating that individuals obtained better cognitive scores after the CCT.

## Discussion

This before-and-after study constitutes the first piece of evidence suggesting that a home-based digital therapeutic program could ameliorate cognitive dysfunction in PASC. The results align with evidence from multiple other conditions involving cognitive dysfunction showing that a CCT yields transfer gains in cognition that can be generalized over time (Hardy et al., 2015).

In addition, this study confirms the findings of previous studies in terms of the severity of the cognitive dysfunction associated with Long COVID, across cognitive domains such as memory, attention, reasoning or coordination (Graham et al., 2021; Jaywant et al., 2021; Woo et al., 2020). The study also provides evidence of the ability of long-haulers to use a digital cognitive evaluation assessment tool to generate a unified general snapshot of their own cognitive status at home. Still, the findings should be taken with the necessary caution, given the methodological limitations associated with a feasibility study, particularly one based before-and-after comparisons, with an unknown denominator of potentially eligible participants and without a control group.

Nevertheless, the favorable direction, frequency and magnitude of the beneficial effects obtained by the participants in this study, warrant more rigorous efforts to determine whether a CCT can in fact improve cognitive functions in COVID long-haulers. In particular, such efforts should be conducted under controlled conditions, and include design features to establish the optimal intensity and duration of the intervention, and whether the effects are sustained.

If confirmed, the findings of this study could open the door for non-invasive, non-pharmacological interventions to curb the cognitive dysfunction that is disabling millions of people in the aftermath of the COVID-19 pandemic.

## Data Availability

All data produced in the present study are available upon reasonable request to the authors

## Acknowledgements

The authors are grateful to the members of the Long Covid ACTS association for their support with participant recruitment, and to Cognifit Inc. for providing access to the platform, the CAB and its normative dataset free of charge.

## Contributions

All authors contributed to the development of the study concept and design. JAD, TJ-G and ARJ contributed to study design, data analysis and interpretation of results. JAD produced the first draft of the manuscript and TJ-G and ARJ contributed to its critical revision and refinement. FM and OB were responsible for participants’ enrollment. All authors approved the final version of the manuscript before submission. Each author contributed important intellectual content during manuscript drafting or revision.

---

**Table 1.** Averaged scores and percentiles in each cognitive domain before and after the CCT

| Cognitive Domain | Pre-test |  |  | Post-test |  |  | Participants that improved | Mean percentile increase | Percentage of improvement |
| --- | --- | --- | --- | --- | --- | --- | --- | --- | --- |
|  | z-score | Percentile | Score 0-800 | z-score | Percentile | Score 0-800 |  |  |  |
| Attention | -0.75 (0.96) | 45 (18) | 361 (148) | -0.11 (0.64) | 59 (18) | 472 (141) | 59/73 | 14 | 31% |
| Memory | -0.22 (0.80) | 48 (21) | 384 (170) | 0.33 (0.81) | 66 (23) | 525 (185) | 63/73 | 18 | 37% |
| Coordination | -1.46 (1.46) | 35 (21) | 280 (167) | -0.45 (1.08) | 53 (24) | 426 (192) | 60/73 | 18 | 52% |
| Perception | -0.66 (0.75) | 42 (14) | 332 (115) | -0.04 (0.70) | 59 (18) | 471 (146) | 64/73 | 17 | 42% |
| Reasoning | -0.56 (0.87) | 44 (19) | 348 (149) | -0.20 (0.77) | 55 (21) | 440 (169) | 56/73 | 11 | 26% |

## References

Bahar-Fuchs A, Barendse MEA, Bloom R, Ravona-Springer R, Heymann A, Dabush H, Bar L, Slater-Barkan S, Rassovsky Y, Schnaider Beeri M. Computerized Cognitive Training for Older Adults at Higher Dementia Risk due to Diabetes: Findings From a Randomized Controlled Trial. J Gerontol A Biol Sci Med Sci. 2020 Mar 9;75(4):747–754. doi: 10.1093/gerona/glz073

Bodner KA, Goldberg TE, Devanand DP, Doraiswamy PM. Advancing Computerized Cognitive Training for MCI and Alzheimer’s Disease in a Pandemic and Post-pandemic World. Front Psychiatry. 2020 Nov 25;11:557571. doi: 10.3389/fpsyt.2020.557571

Buades-Sitjar, F., Duñabeitia, J.A. Intelligence subcomponents and their relationship to general knowledge. J Cult Cogn Sci (2022). https://doi.org/10.1007/s41809-022-00113-z

Bungenberg J, Humkamp K, Hohenfeld C, Rust MI, Ermis U, Dreher M, Hartmann NK, Marx G, Binkofski F, Finke C, Schulz JB, Costa AS, Reetz K. Long COVID-19: Objectifying most self-reported neurological symptoms. Ann Clin Transl Neurol. 2022 Feb;9(2):141–154. doi: 10.1002/acn3.51496.

Centers for Disease Control and Prevention. Long COVID or Post-COVID Conditions. 2022. https://www.cdc.gov/coronavirus/2019-ncov/long-term-effects/index.html

Davis HE, Assaf GS, McCorkell L, Wei H, Low RJ, Re’em Y, Redfield S, Austin JP, Akrami A. Characterizing long COVID in an international cohort: 7 months of symptoms and their impact. EClinicalMedicine. 2021 Aug;38:101019. doi: 10.1016/j.eclinm.2021.101019

Deer RR, Rock MA, Vasilevsky N, Carmody L, Rando H, Anzalone AJ, Basson MD, Bennett TD, Bergquist T, Boudreau EA, Bramante CT, Byrd JB, Callahan TJ, Chan LE, Chu H, Chute CG, Coleman BD, Davis HE, Gagnier J, Greene CS, Hillegass WB, Kavuluru R, Kimble WD, Koraishy FM, Köhler S, Liang C, Liu F, Liu H, Madhira V, Madlock-Brown CR, Matentzoglu N, Mazzotti DR, McMurry JA, McNair DS, Moffitt RA, Monteith TS, Parker AM, Perry MA, Pfaff E, Reese JT, Saltz J, Schuff RA, Solomonides AE, Solway J, Spratt H, Stein GS, Sule AA, Topaloglu U, Vavougios GD, Wang L, Haendel MA, Robinson PN. Characterizing Long COVID: Deep Phenotype of a Complex Condition. EBioMedicine. 2021 Dec;74:103722. doi: 10.1016/j.ebiom.2021.103722

Embon-Magal S, Krasovsky T, Doron I, Asraf K, Haimov I, Gil E, Agmon M. The effect of co-dependent (thinking in motion [TIM]) versus single-modality (CogniFit) interventions on cognition and gait among community-dwelling older adults with cognitive impairment: a randomized controlled study. BMC Geriatr. 2022 Aug 31;22(1):720. doi: 10.1186/s12877-022-03403-x

Gavelin HM, Domellöf ME, Leung I, Neely AS, Launder NH, Nategh L, Finke C, Lampit A. Computerized cognitive training in Parkinson’s disease: A systematic review and meta-analysis. Ageing Res Rev. 2022 Sep;80:101671. doi: 10.1016/j.arr.2022.101671

Graham EL, Clark JR, Orban ZS, Lim PH, Szymanski AL, Taylor C, DiBiase RM, Jia DT, Balabanov R, Ho SU, Batra A, Liotta EM, Koralnik IJ. Persistent neurologic symptoms and cognitive dysfunction in non-hospitalized Covid-19 “long haulers”. Ann Clin Transl Neurol. 2021 May;8(5):1073–1085. doi: 10.1002/acn3.51350..” ACTN, 8(5), 1073–1085. https://doi.org/10.1002/acn3.51350

Hardy JL, Nelson RA, Thomason ME, Sternberg DA, Katovich K, Farzin F, Scanlon M. Enhancing Cognitive Abilities with Comprehensive Training: A Large, Online, Randomized, Active-Controlled Trial. PLoS One. 2015 Sep 2;10(9):e0134467. doi: 10.1371/journal.pone.0134467

Islam MF, Cotler J, Jason LA. Post-viral fatigue and COVID-19: lessons from past epidemics. Fatigue: Biomed. Health Behav, 2020; 8(2), 61–69. doi: 10.1080/21641846.2020.1778227

Jaywant A, Vanderlind WM, Alexopoulos GS, Fridman CB, Perlis RH, Gunning FM. Frequency and profile of objective cognitive deficits in hospitalized patients recovering from COVID-19. Neuropsychopharmacology. 2021 Dec;46(13):2235–2240. doi: 10.1038/s41386-021-00978-8

Lancaster GA, Thabane L. Guidelines for reporting non-randomised pilot and feasibility studies. Pilot Feasibility Stud. 2019 Oct 6;5:114. doi: 10.1186/s40814-019-0499-1

Ledford H. Long-COVID treatments: why the world is still waiting. Nature. 2022 Aug;608(7922):258–260. doi: 10.1038/d41586-022-02140-w

Moghimi N, Di Napoli M, Biller J, Siegler JE, Shekhar R, McCullough LD, Harkins MS, Hong E, Alaouieh DA, Mansueto G, Divani AA. The Neurological Manifestations of Post-Acute Sequelae of SARS-CoV-2 infection. Curr Neurol Neurosci Rep. 2021 Jun 28;21(9):44. doi: 10.1007/s11910-021-01130-1

National Institute for Health and Care Excellence. COVID-19 rapid guideline: managing the long-term effects of COVID-19. 2021. https://www.nice.org.uk/guidance/NG188

Premraj L, Kannapadi NV, Briggs J, Seal SM, Battaglini D, Fanning J, Suen J, Robba C, Fraser J, Cho SM. Mid and long-term neurological and neuropsychiatric manifestations of post-COVID-19 syndrome: A meta-analysis. J Neurol Sci. 2022 Mar 15;434:120162. doi: 10.1016/j.jns.2022.120162

Reuschke D, Houston D. The impact of Long COVID on the UK workforce, Appl Econ Lett. 2022. doi: 10.1080/13504851.2022.2098239

Soriano JB, Murthy S, Marshall JC, Relan P, Diaz JV; WHO Clinical Case Definition Working Group on Post-COVID-19 Condition. A clinical case definition of post-COVID-19 condition by a Delphi consensus. Lancet Infect Dis. 2022 Apr;22(4):e102–e107. doi: 10.1016/S1473-3099(21)00703-9

Stefano GB. Historical Insight into Infections and Disorders Associated with Neurological and Psychiatric Sequelae Similar to Long COVID. Med Sci Monit. 2021 Feb 26;27:e931447. doi: 10.12659/MSM.931447

Taquet M, Dercon Q, Luciano S, Geddes JR, Husain M, Harrison PJ. Incidence, co-occurrence, and evolution of long-COVID features: A 6-month retrospective cohort study of 273,618 survivors of COVID-19. PLoS Med. 2021 Sep 28;18(9):e1003773. doi: 10.1371/journal.pmed.1003773

The jamovi project (2021). jamovi (Version 1.6) [Computer Software]. Retrieved from https://www.jamovi.org

Woo MS, Malsy J, Pöttgen J, Seddiq Zai S, Ufer F, Hadjilaou A, Schmiedel S, Addo MM, Gerloff C, Heesen C, Schulze Zur Wiesch J, Friese MA. Frequent neurocognitive deficits after recovery from mild COVID-19. Brain Commun. 2020 Nov 23;2(2):fcaa205. doi: 10.1093/braincomms/fcaa205

World Health Organization. In the wake of the pandemic Preparing for Long COVID. 2021. https://apps.who.int/iris/bitstream/handle/10665/339629/Policy-brief-39-1997-8073-eng.pdf

Xu, E., Xie, Y. & Al-Aly, Z. Long-term neurologic outcomes of COVID-19. Nat Med (2022). https://doi.org/10.1038/s41591-022-02001-z

Yaneva A, Massaldjieva R, Mateva N. Initial Adaptation of the General Cognitive Assessment Battery by Cognifit™ for Bulgarian Older Adults. Exp Aging Res. 2022 Jul-Sep;48(4):336–350. doi: 10.1080/0361073X.2021.1981096

Zhou Y, Feng H, Li G, Xu C, Wu Y, Li H. Efficacy of computerized cognitive training on improving cognitive functions of stroke patients: A systematic review and meta-analysis of randomized controlled trials. Int J Nurs Pract. 2022 Jun;28(3):e12966. doi: 10.1111/ijn.12966

